# Role of Blood Lipids in the Shared Genetic Etiology Between Major Depressive Disorder and Myocardial Infarction: A Large-scale Multi-trait Association Analysis

**DOI:** 10.1101/2024.04.08.24305481

**Authors:** Yanchen Zhu, Zhengbo Wu, Yahui Wang, Zhaorui Cui, Fani Liu, Jiqiang Hu

**Affiliations:** Cardiology Department, Dongfang Hospital Beijing University of Chinese Medicine, Beijing, China

**Keywords:** Major depressive disorder, Myocardial infarction, Blood lipids, Shared genetics, Multi-trait association analysis

## Abstract

**Background:** The genetic role of blood lipids (BL) in the shared genetic etiology between major depressive disorder (MDD) and myocardial infarction (MI) has not been fully characterized.

**Methods:** We first evaluated genetic associations and causal inferences between MDD, MI and the quantitative traits of BL. To further unravel the underlying genetic mechanisms, we performed multi-trait association analysis to identify novel or pleiotropic genomic risk loci, and shared causal variants for diseases involving BL. Using multiple post-GWAS methods, we explored potential genes, pathways, tissues, cells, and therapeutic targets associated with diseases from different perspectives.

**Findings:** We found extensive global and local genetic correlations between MDD, MI and the traits of BL. Mendelian randomization (MR) analyses showed that lipid metabolism mediated 26.5% of the mediating effect of MDD leading to MI. Multi-trait association analysis successfully identified 13 MDD- and 36 MI- novel risk loci which have never been reported before. Notably, many pleiotropic loci and shared causal variants were identified across risk loci for both diseases, such as 11q23.3 (rs117937125) and 12q13.3 (rs188571756), which also colocalized for traits of BL. Pathway enrichment analysis further highlighted shared biological pathways primarily involving synaptic function, arterial development, and lipid metabolism. Lastly, gene-mapping, gene-based, transcriptome-wide and proteome-wide association, and MR-proteomic analyses revealed candidate pathogenic genes and therapeutic targets (such as ANGPTL4 and TMEM106B).

**Interpretation:** These findings not only provide novel insights into the role of BL in the comorbidity between MDD and MI, but also benefit the development of preventive or therapeutic drugs for diseases.

## Introduction

Major depressive disorder (MDD) and myocardial infarction (MI) are universal leading causes of disability worldwide^1^. The epidemiological observations have repeatedly demonstrated that these two diseases are highly comorbid^2,3^. However, the underlying mechanism between MDD and MI remains unclear. Abnormal blood lipids (BL) are established risk factors for MI, and MDD may lead to dyslipidemia^4,5^. Consequently, BL may play a significant role in the comorbidity of MDD and MI. Further exploration of their interplay could elucidate important disease pathways and interventions with added clinical benefit.

Previous genome-wide association studies (GWAS) have identified several potential shared genomic loci affecting BL, MDD and MI^6,7^, providing further evidence of common biological pathways between these phenotypes. Using genetic variants as instrumental variables can infer the genetic role of BL in the relationship between MDD and MI. Multi-trait joint analysis can further unravel the genetic mechanisms underlying their comorbidities. Multi-trait analysis of GWAS (MTAG) enabled researchers effectively to increase the sample size from multiple related traits and has become an effective statistical method to improve statistical power to identify novel genomic risk loci for target traits^8^. The latest genetic association study of quantitative traits of BL in more than 1 million people showed significant polygenic heritability and numerous genomic loci^9^. Therefore, their multi-trait joint analysis with MDD and MI can not only borrow information from these traits to deeply explore disease-related genetic variations, but also identify pleiotropic loci shared by BL, MDD, and MI.

In this study, using largest publicly available GWAS summary statistics, we evaluated genetic associations and causal inferences for BL, MDD, and MI, and then performed multi-trait analysis between these phenotypes. Our study aims to achieve two primary objectives. Firstly, we aim to discover novel or pleiotropic genomic risk loci, and shared causal variants for diseases involving BL, elucidate their genetic mechanism, and explore potential drug targets. Secondly, we aim to characterize the genetic roles of BL in MDD and MI, as well as investigate their shared genetic etiology basis.

## Methods

### 1. Population samples and ethics

We utilized the most recent and largest publicly available GWAS summary statistics from European ancestry individuals in this study. GWAS for MDD were obtained from the Psychiatric Genomics Consortium (PGC)^10^, which meta-analyzed 33 cohorts with 170,756 cases and 329,443 controls. GWAS for MI were obtained from a meta-analysis study among 639,221 participants (61,505 cases and 577,716 controls) from UK Biobank and CARDIoGRAMplusC4D consortium^11^. The Global Lipids Genetics Consortium (GLGC) consortium provided summary statistics for the quantitative traits of BL [high-density lipoprotein cholesterol (HDL-C), low-density lipoprotein cholesterol (LDL-C), total cholesterol (TC), and triglyceride (TG)] (N= 1,320,016)^9^. The Supplementary Table 1 provides access to the specific details of each GWAS summary statistics.

### 2. Statistical analysis

Fig. 1 presents a schematic overview of our study. All GWAS summary statistics underwent genotypic quality control measures. Supplement Methods provide details of these methods.

**Fig.1.**
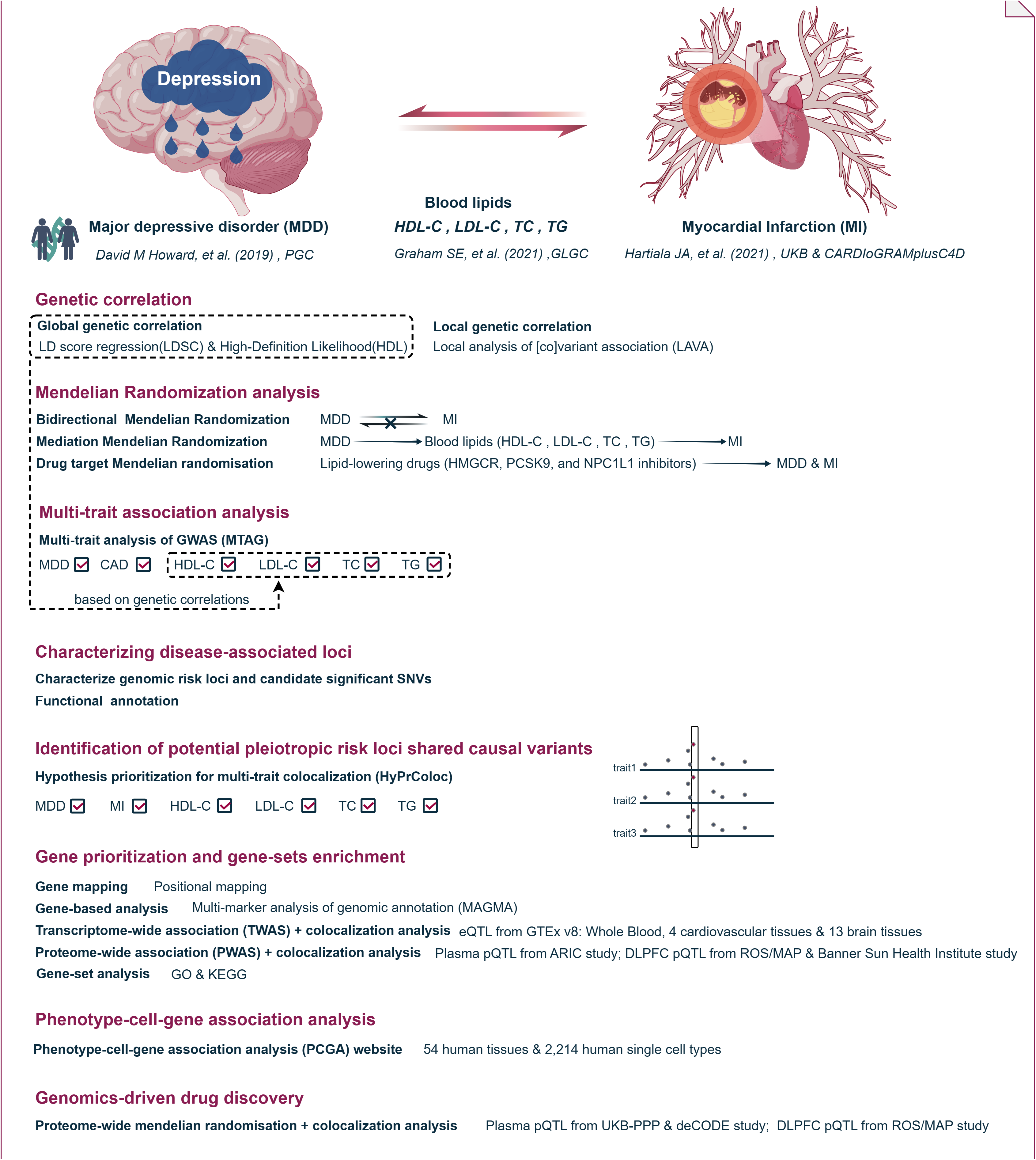
Design schematic of the present study. MDD: major depressive disorder, MI: myocardial infarction, HDL-C: high-density lipoprotein cholesterol, LDL-C: low-density lipoprotein cholesterol, TC: total cholesterol, TG: triglyceride.

We used both linkage disequilibrium (LD) score regression (LDSC)^12^ and high-definition likelihood (HDL)^13^ to evaluate heritability of each trait and the genetic correlation between MDD, MI, and the quantitative traits of BL. Given the intricate genetic structure of each region, local analysis of [co]variant association (LAVA)^14^ was used to assess local genetic correlation. Next, we used Mendelian randomization (MR)^15^ to explore potential causal associations and mediating effects between them. Using genetic proxies for drug target genes, we further explored the effects of lipid-lowering drugs on MDD. For the above statistical analyses, we applied the Benjamini-Hochberg false discovery rate (FDR) approach with a threshold of 0.05 to correct for multiple testing.

To further unravel the genetic mechanisms underlying the comorbidity, we utilized MTAG^8^ to identify potential pleiotropic SNVs through the interrelationships of pairwise traits with significant genetic correlation. We calculated the maximum false discovery rate (maxFDR) to evaluate the overall inflation caused by the violation of the assumption of homogeneity in MTAG analysis. The Functional Mapping and Annotation of Genetic Associations (FUMA)^16^ was applied to further characterize potential pleiotropic loci. By focusing on consecutive non-overlapping genetic risk loci for MDD and MI, we used hypothesis prioritization in multi-trait colocalization (HyPrColoc)^17^ analysis to identify shared causal variants within each genomic locus across traits. We considered a colocalized locus with the posterior probability of H4 (PP.H4) exceeded 0.75.

Based on MTAG results, we further explored the shared biological mechanisms of these pleiotropic loci. Using the GO and KEGG databases, we conducted gene-set enrichment analyses to identify biological pathways. Using DESE (driver tissue estimation by selective expression)^18^, we explored the tissue/cell types specificity associated with diseases through the phenotype-cell-gene association analysis (PCGA)^18–21^ website.

To explore candidate pathogenic genes for MDD and MI in MTAG, we performed a combination of gene-mapping and multi-marker analysis of genomic annotation (MAGMA)^22^ to identify potential candidate genes (at FDR < 0.05). Next, we used transcriptome-wide association studies (TWAS)^23^ and proteome-wide association studies (PWAS)^24^ in combination with bayesian colocalization^25^ (PP.H4>0.75) to generate hypotheses of target genes and directions of effects. We restricted the analysis to cardiovascular and brain tissues and considered FDR < 0.05 as the threshold of significant associations. We used precomputed cross-tissue weights from eQTL reference panels in GTEx v.8 (across 5 cardiovascular and 13 brain tissues), as well as the weights for protein expression in plasma (ARIC study)^26^ and in dorsolateral prefrontal cortex (DLPFC) (ROS/MAP study^27^ and the Banner Sun Health Institute study^28^). To further identify therapeutic targets and enhance gene-driven drug discovery, we utilized proteome-wide MR^15^ (FDR < 0.05) and Bayesian colocalization^25^ (PP.H4 > 0.75) analysis to identify potential causal therapeutic plasma [UK Biobank Pharma Proteomics Project (UKB-PPP)^29^ and deCODE genetics^30^] and brain (ROS/MAP study^27^) pleiotropic and specific protein targets for MDD and MI.

### 3. Genomic Loci Characterization and Functional Annotation

For significant pleiotropic single nucleotide variants (SNVs) from MTAG, we used the FUMA to characterize significant genomic loci (P < 5×10^−8^). Using LD information from the 1000 Genome Project phase 3 reference panel of European population, FUMA identified genome-wide significant independent SNVs with r^2^ less than 0.6 and lead SNVs with r^2^ less than 0.1 within 1 Mb. Risk loci were defined by combining lead SNVs that physically overlapped or had LD blocks within 250 kb apart. FUMA additionally provided functional annotations such as ANNOVAR analysis, combined annotation dependent depletion (CADD) scores, and RegulomeDB scores. Variants with CADD score exceeding 12.37 were deemed potentially deleterious.

## Results

### Genetic correlation and genetic causal inference

Both LDSC and HDL results indicated that MDD demonstrated significant positive genetic correlations with MI (rg_LDSC_ = 0.19 and rg_HDL_ = 0.23) (Fig.2; Supplementary Table 2-3). LDL-C, TC and TG exhibited significant positive genetic correlations with both MDD and MI. Conversely, HDL-C displayed significant negative genetic correlations. LAVA discovered a total of significant bivariate local genetic correlations for MDD with MI at 28 specific regions (FDR.P<0.05) (Fig.2; Supplementary Table 4). And we also found a total of 198, 505 specific regions of significant local genetic correlations (FDR.P<0.05) for quantitative traits of BL with MDD and MI, respectively. The coexistence of both negative and positive local genetic correlations suggests a multifaceted impact. The tendency of overall positive and negative local correlations aligned with the direction of their respective global correlations.

**Fig.2.**
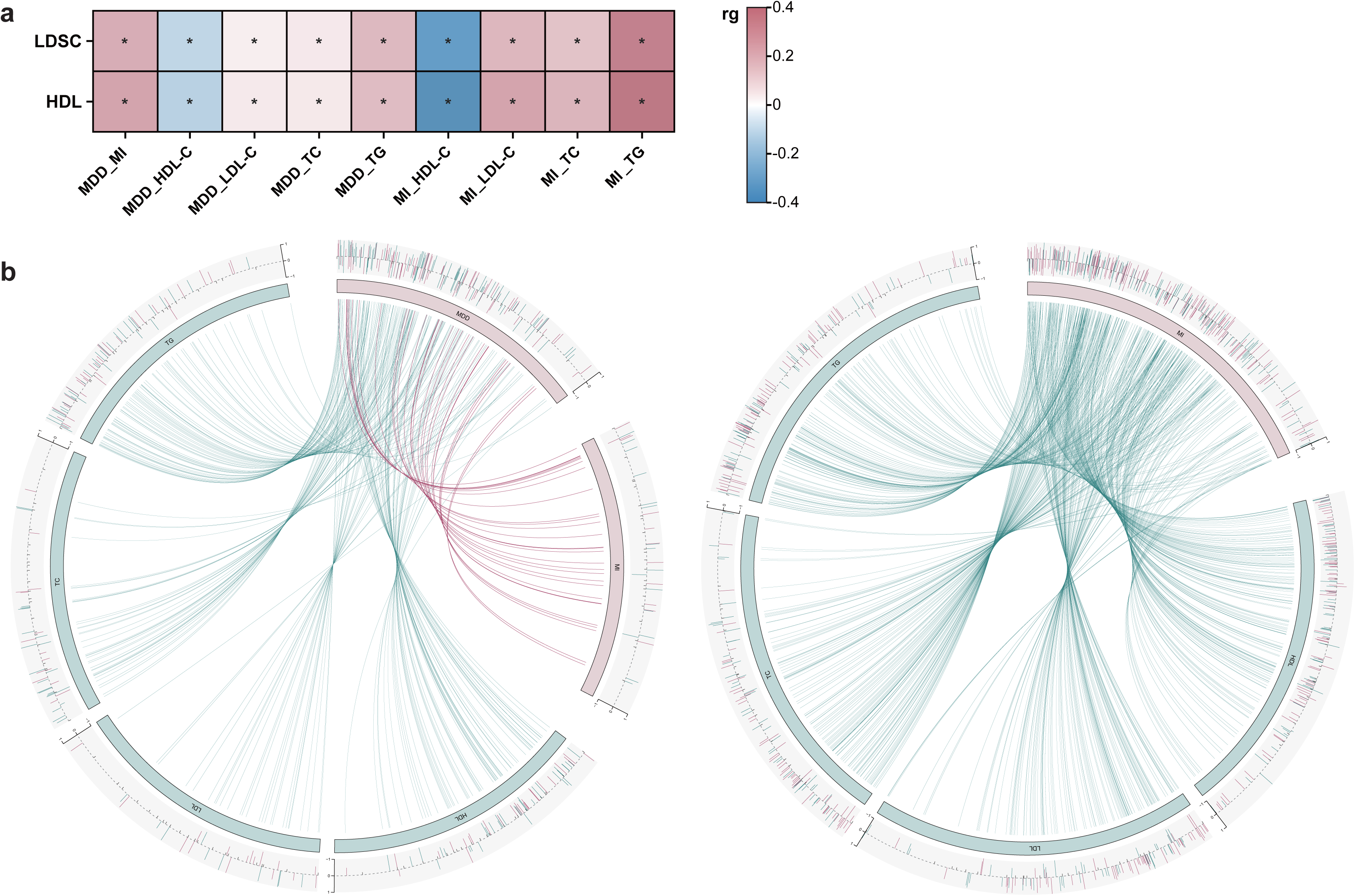
Genetic correlations between major depressive disorder, myocardial infarction, and the quantitative traits of blood lipids. a, the correlation heat-map shows the global genetic correlations between major depressive disorder, myocardial infarction and the quantitative traits of blood lipids. b, chord diagram shows their local genetic correlations. See Supplementary Table 2-4 for complete details of genetic correlations estimates. *LDSC: linkage disequilibrium score regression, HDL: high-Definition Likelihood*.

MR analysis revealed that MDD increased the risk of MI unidirectionally (or = 1.23, se = 0.04; P = 8.63×10^-06^) (Fig.3; Supplementary Table 5). Mediation analysis further indicated that LDL-C, TC, and TG mediated this effect, accounting for 7.5%, 5.7%, and 13.3% of the total effect, respectively (Fig.3; Supplementary Table 5). Subsequent drug-target MR (Supplement Methods) analysis demonstrated that while commonly prescribed lipid-lowering medications (HMGCR, PCSK9, and NPC1L1 inhibitors) effectively reduced the risk of MI, HMGCR and PCSK9 inhibitors were associated with an increased risk of MDD (Fig.3). Conversely, there was no discernible effect of NPC1L1 inhibitors on MDD.

**Fig.3.**
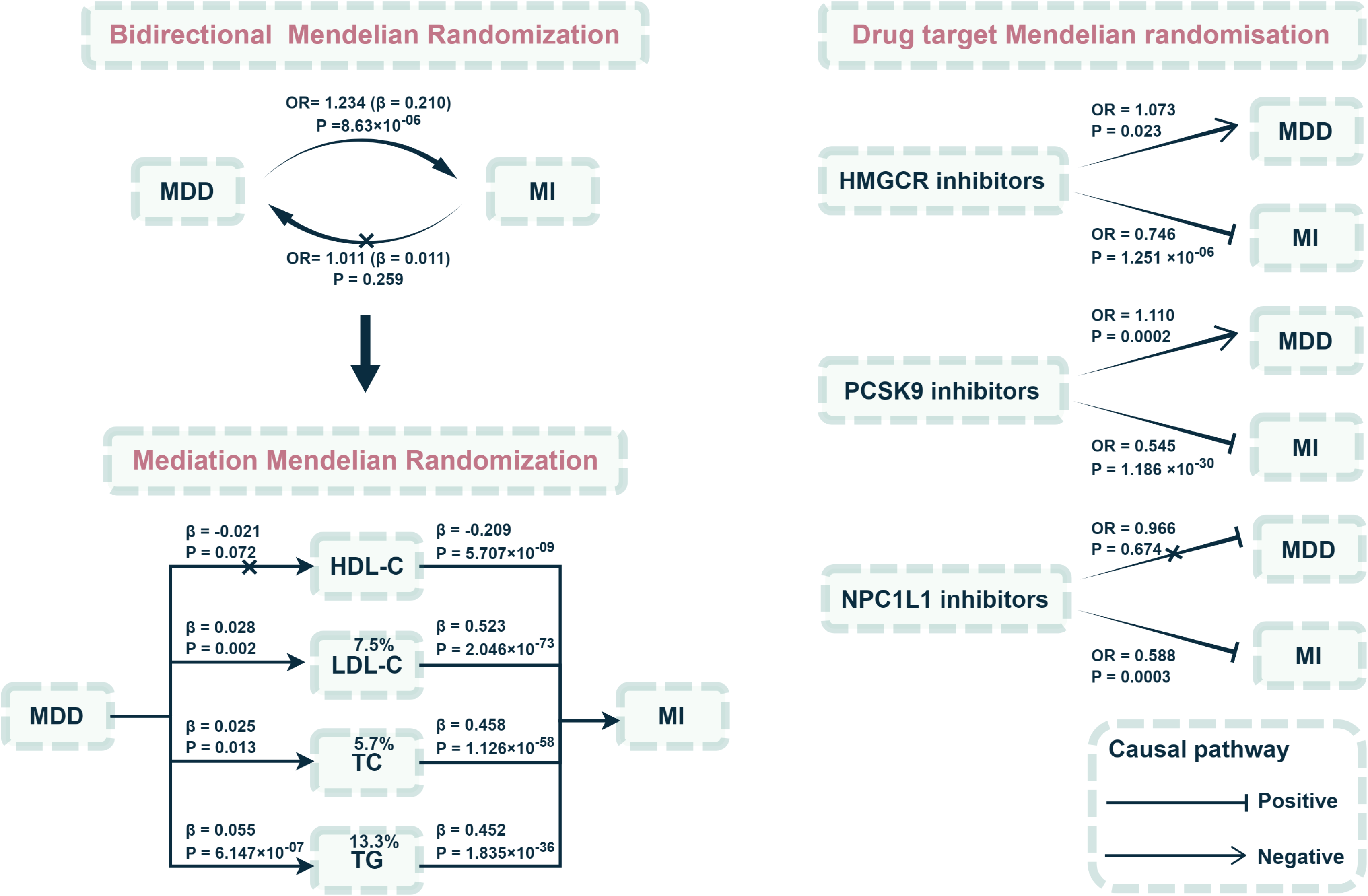
Summary of the causal evidence from mendelian randomization analyses. See Supplementary Table 5 for complete details of Mediation Mendelian randomization results. *MDD: major depressive disorder, MI: myocardial infarction, HDL-C: high-density lipoprotein cholesterol, LDL-C: low-density lipoprotein cholesterol, TC: total cholesterol, TG: triglyceride*.

### Multi-trait association analysis and functional annotation

We performed a meta-analysis of -GWAS_MDD_, -GWAS_MI_, and -GWAS_all traits of BL_ using MTAG. MTAG analysis significantly increased the effective sample sizes and heritability of diseases. The polygenic heritability exhibited an impressive 1.04-, and 1.28-fold escalation for MDD and MI consideration (Fig.4). We calculated max-FDR values of 0.0006, and 0.0018 for MDD and MI, respectively, suggesting no overall inflation due to violation of the homogeneous assumption. We successfully identified 63 and 120 genomic risk loci from for MDD, and MI, of which 19 and 54 are novel (Supplementary Table 6-7). By comparing loci reported in GWAS catalog (Supplementary Table 8-9), we found that a total of 13 MDD- and 36 MI- loci which have not been reported previously. Multi-trait colocalization analysis from HyPrColoc highlighted 32 pleiotropic loci on the consecutive non-overlapping genetic risk loci for MDD and MI, which were colocalized to share a causal variant (Supplementary Table 10). Five of 32 pleiotropic loci were identified to be colocalized among MDD, MI and the quantitative traits of BL, including 1q41 (rs142709330), 11q23.3 (rs117937125), 12q13.3 (rs188571756), 16p13.11 (rs112527105), 19p13.11 (rs145793672) locus. These findings supported the significant role of BL in the comorbidity.

**Fig.4.**
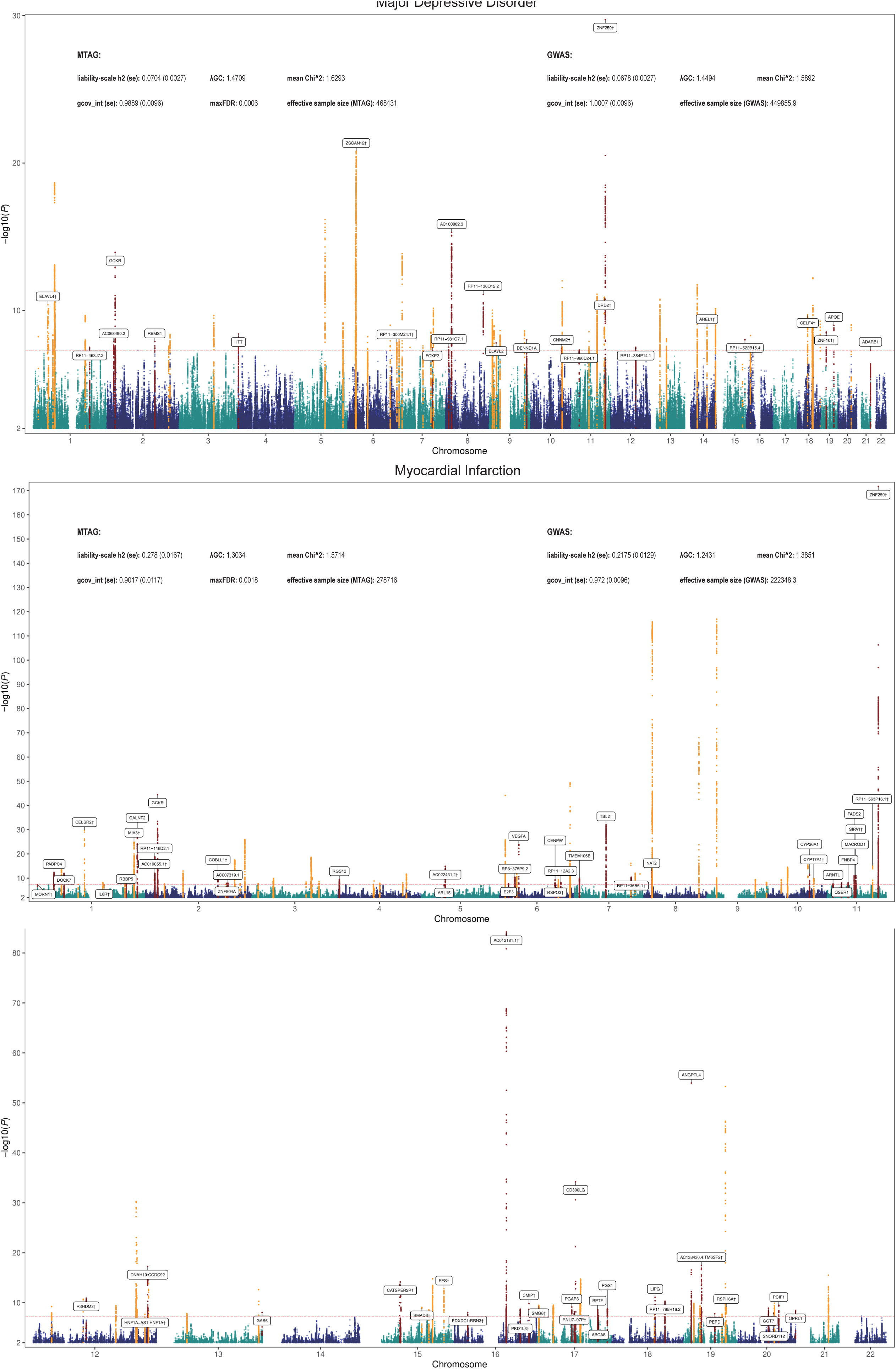
Manhattan plots of MTAG results for major depressive disorder and myocardial infarction. The red line marks the genome-wide significance threshold (5 × 10^−8^). Novel genomic risk loci are highlighted in red, and the loci previously reported in GWAS are in orange. Annotated genes include only these nearest genes to the top signals from novel and colocalized loci. See Supplementary Table 6-7 and 10 for complete details of genomic risk loci. *†:colocalized loci, h2: heritability (LD score regression), se: standard error, gcov_int: genetic covariance intercept,* λ*GC: the genomic control inflation factor based on the median, maxFDR: maximum FDR*.

Functional annotation of the 202 independent significant SNVs across 63 MDD risk loci and 652 SNVs from MI loci revealed that most variants were intergenic or intronic (Supplementary Table 11-12). For MDD, six (2.97%) of 202 MDD SNVs were exonic variants, including 4 messenger RNA (mRNA) exonic variants (1 synonymous SNVs, 3 nonsynonymous SNVs) and 2 noncoding RNA exonic variants. The index SNV rs2232423 was the most significant mRNA exonic variant (P_MTAG_ = 1.59×10^-21^) at 6p22.2 locus, regulating the expression of ZSCAN12. ZSCAN12 has been identified as a pivotal gene associated with depression and schizophrenia, exhibiting widespread expression in brain tissue^31,32^. For MI, we identified 28 (4.29%) exonic variants from 652 SNVs, of which 21 were missense variants (3 synonymous SNVs, 18 nonsynonymous SNVs). The most significant exonic variant of coding RNA was rs116843064 (P_MTAG_ = 1.07×10^-54^, gene: ANGPTL4) at 19p13.2 locus, followed by rs268 (P_MTAG_ = 2.10×10^-54^, gene: LPL) at 8p21.3 locus, and rs429358 (P_MTAG_ = 5.30 ×10^-54^, gene: APOE) at 19q13.32 locus. Further annotation by CADD scores predicted that 14 MDD- and 45 MI- SNVs were deleterious (CADD score > 12.37). The variant with the highest CADD score was also the exonic variant rs116843064 (CADD score 33 for MI), which encoding ANGPTL4 protein. ANGPTL4 can mediate inactivation of the lipoprotein lipase LPL, therefore playing an important role in the regulation of triglyceride clearance of lipid metabolism^33^.

### Gene-set enrichment

MAGMA gene-set enrichment found 19 MDD- and 78 MI- significantly enriched pathways (FDR < 0.05) (Supplementary Fig.1). These analyzes showed that MDD is significantly associated with biological processes involving various functions in the synapse, such as branching morphogenesis of nerves and synaptic assembly. In addition, this analysis revealed the association of MDD with pathways related to lipid metabolism, particularly with the biological processes of triglyceride-rich lipoprotein remnant particles and phosphatidylcholine-sterol O-acyltransferase (LCAT) activator activity molecular function. Gene-sets involved with this activity include important lipid-regulated genes such as APOA1, APOC1, and APOE. Of which, APOE was located at the novel MDD locus 19q13.32. MI was significantly associated with biological processes or molecular functions related to lipid metabolism and arterial development, such as artery development and reactome plasma lipoprotein remodeling. Notably, MI also involves the molecular function of LCAT activator activity. Within the above gene-sets, angiopoietin-like protein 4 (ANGPTL4) is the key gene, implicated at the novel risk locus 19p13.2 of MI. Lipisense, an ANGPTL4 inhibitor, is a novel drug for the treatment of severe hypertriglyceridemia. Over all, these results demonstrate the important role of lipid metabolism in both diseases.

### Tissue and cell type enrichment specificity association analysis

To further illuminate the underlying biological causes of diseases, we investigated tissue/cell type specificity associated with diseases based on DESE (Supplement Methods). In 54 human tissues, we found that MDD was significantly associated with brain and pituitary tissues (Supplementary Fig. 2). These results highlighted almost all brain regions, including the cerebral cortex and subcortical structures such as the hypothalamus, hippocampus, and amygdala. The role of the pituitary in MDD may reflect the potential effects of hormones on cardiac metabolism. MI was primarily associated with heart and adipose tissues, such as coronary and visceral adipose. Using the 2,214 types of human cells reference dataset for cell type-specific analysis showed that MDD was predominantly associated with excitatory and inhibitory neurons, and other brain cells such as astrocytes, oligodendrocytes or microglial cells (Supplementary Fig. 3). MI was significantly enriched in cell types involving macrophages, endothelial cells, smooth muscle cells, and fibroblasts, which may reflect important processes in the inflammatory response, tissue repair, regulation of vascular function, and lipid homeostasis.

### Gene-based analyses and gene prioritization

FUMA mapped 155 and 426 positional mapped genes for MDD and MI (within 0kb from the locus) (Supplementary Table 13-14). Based on the mapping genes, the MAGMA gene-based analysis found a total of 149 MDD, and 499 MI genes (FDR.P<0.05) (Supplementary Table 15-16).

Using TWAS-Fusion and eQTL based on tissue-specific eQTL data (GTEx v8) form 5 cardiovascular tissues and 13 brain tissues, we identified 146 and 292 genes at the transcriptome-wide level (FDR.P<0.05) and colocalized (PP. H4>0.75) in at least one tissue for MDD and MI, respectively (Supplementary Table 17-18). 119 functional genes targeting MDD were mapped outside of the MDD-risk loci, and 198 MI genes were mapped outside of the MI-risk loci. Furthermore, using similar parameters in TWAS-Fusion, we conducted PWAS analyses with pQTL from plasma [ARIC study (n = 7,213 European Americans, n = 4,657 proteins)] and DLPFC tissues [ROS/MAP study (n = 376 individuals, n = 1,475 proteins) and Banner Sun Health Institute study (n = 152 individuals, n = 1,145 proteins)]. PWAS analyses revealed 19 MDD and 46 MI genes with colocalization evidence (Supplementary Table 19-20). Overall, we observed extensive functional genes targeting both MDD and MI in brain and cardiovascular tissues and these genes have the same direction of effect on diseases in different tissues, which potentially revealed the importance of the heart-brain axis in MDD and MI. In TWAS and PWAS, 33 pleiotropic transcript or protein abundances were associated with the risk of both MDD and MI.

### Proteome-wide plasma and brain therapeutic targets

We used cis-pQTL as the genetic instrument for MR analysis to systematically assess the evidence for causal effects of plasma and DLPFC proteins on MDD and MI. Combining two-sample MR (FDR. P<0.05) and colocalization (PP. H4>0.75) analyses, a total of 22 MDD and 68 MI protein target genes were successfully identified in at least one dataset (Fig.5; Supplementary Table 21-22). Sensitivity analyses indicated no reverse causation (steiger test) and no horizontal pleiotropy (MR- Egger regression intercept test). Four target genes (MDD: PSMB4 and TMEM106B; MI: ACP1 and ALDH2) reached significant levels in both plasma and DLPFC tissues. In addition, we identified eleven pleiotropic proteins (ANGPTL4, APOA1, APOC3, ATP13A1, CNNM2, DUSP13, GCKR, GPN1, NRBP1, TMEM106B, and ZPR1) targeting MDD and MI. Notably, these proteins had effects in the same direction on both MDD and MI, which are expected to be potential therapeutic targets for the comorbidities.

**Fig.5.**
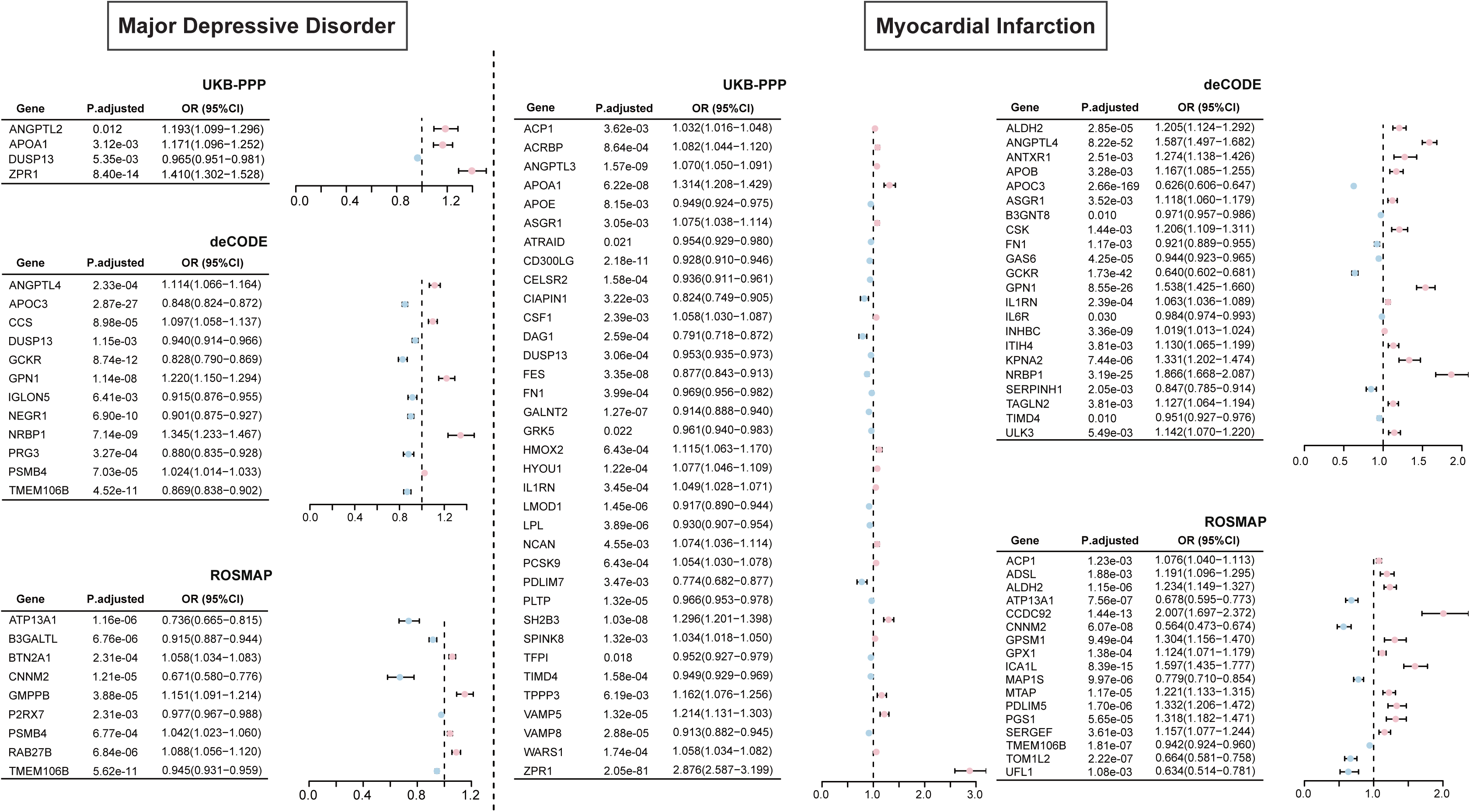
Forest plots of protein targets for major depressive disorder and myocardial infarction indicated by proteome-wide Mendelian randomization analyses. The forest plot displayed 22 MDD- and 68 MI- protein targets that satisfied the statistical significance of both Mendelian randomization (FDR.P<0.05) and colocalization (PP.H4>0.7) in at least one cohort. The square symbolizes the odds ratio, the horizontal line represents the 95% confidence interval. See Supplementary Table 21-22 for complete details of the proteome-wide Mendelian randomization results. *MDD: major depressive disorder, MI: myocardial infarction, UKB-PPP: UK Biobank Pharma Proteomics Project; deCODE: deCODE genetics, ROSMAP: Religious Orders Study/Memory and Aging Project*.

## Discussion

The intertwined connections between heart and brain health are under increasing attention, with the metabolism potentially playing an important role in the heart-brain axis^34^. In this study, we found extensive genome-wide genetic correlations and genetic overlap between MDD, MI, and the quantitative traits of BL, and determined their causal roles. MTAG analysis significantly improved the statistical power to identify novel genomic risk loci for the comorbidity and identified 13 MDD- and 36 MI- novel risk loci which have never been reported before. Further comprehensive post-GWAS analyses highlighted the pleiotropic loci, potential shared causal variants, potential genes, pathways, tissues, cells,and therapeutic targets. These findings furthered our understanding of the role of BL in the shared genetic etiology in the co-morbidity between MDD and MI, and provided novel insights for therapeutic intervention.

MR analyses showed that lipid metabolism mediated 26.5% of the mediating effect of MDD leading to MI. Therefore, patients with MDD are required intensive monitoring and/or treatment of lipids. In addition, we found that commonly used lipid-lowering drugs (HMGCR and PCSK9 inhibitors) in turn increased the risk of MDD, which may pose a challenge to the existing pharmacologic treatment of MI and MDD co-morbidities. This situation may also explain part of the potential cause of new-onset MDD in patients with MI^35,36^. However, some previous studies have yielded opposite or negative conclusions, so the effects of lipid-lowering drugs on depression may need to be interpreted with caution^37,38^.

With the investigation of the underlying pleiotropic associations, MTAG analysis deepened our understanding of the genetic mechanisms underlying them. Overall, pleiotropic variants between MDD and MI were extensively distributed, with some loci especially highlighted the important role of lipids. For instance, the 11q23.3 locus, which was identified as novel risk loci for both MDD and MI, was colocalized between MDD, MI, and all quantitative traits of BL, with the same potential shared causal variant rs117937125 identified (mapped gene: ZNF259). Previous studies reported that ZNF259 is associated with dyslipidemia in the whole population and that its transcriptional upregulation increases the sensitivity of coronary endothelial cells to oxidative stress and inflammation^39^. These results highlighted the important role of lipids in the comorbidity. In addition, enrichment analyses have also pointed to the presence of important lipid metabolic pathways in MDD. For example, both MDD and MI involved the molecular function of LCAT activator activity. LCAT is an enzyme that play a critical role in lipid metabolism, and it binds to both HDL and LDL in plasma^40,41^. LCAT activation facilitate removal of excess cholesterol from the arterial wall and prevents the formation of atherosclerotic plaques.

We conducted tissue-specific multi-omics analyses (TWAS, PWAS, MR with pQTL) to provide genetic evidence for pathophysiological mechanisms and therapeutic targets for MDD and MI in both brain and heart. Of note, colocalization analyses confirmed that the above significant associations observed did not represent random correlations between gene expression and non-causal variants. Our findings indicated the existence of widely expressed pleiotropic functional genes in both brain- and heart-related tissues, and these genes have the same effect direction on disease in different tissues. MR-proteomic analyses uncovered the putative causal role of human proteins in MDD and MI, which improves the success of clinical trials for co-morbid drug discovery.

There are limitations acknowledged in this study. Our analyses were restricted to individuals of European ancestry only and the results may not be generalized to other ancestries. We excluded all rare variants (MAF < 1%) from the genotypic quality control and consequently from all following analyses. Therefore, we may not be able to identify rare variants with large effects. In MR analyses, we exclusively utilized cis-regulatory regions as instrumental variables, which may mitigate horizontal pleiotropy to some extent and result in reduced statistical efficacy. Finally, our study was limited to analyzing and validating pQTL data with available instrumental variables, which may overlook some potential therapeutic targets.

## Conclusion

In summary, this multi-trait association study revealed the role of BL in the shared genetic etiology between MDD and MI. We found that pleiotropic genetic variants, loci, and genes were extensively distributed across the genome. We highlighted underlying potential biological mechanisms and therapeutic targets. These findings not only provide novel insights into the shared genetic basis for the co-morbidity between MDD and MI, but also benefit the development of preventive or therapeutic drugs for diseases.

## Contributors

YZ, Conceptualization, Data curation, Formal analysis, Methodology, Writing-original draft; ZW and YW, Conceptualization, Methodology, Writing-original draft; ZC and FL, Conceptualization, Methodology, Writing-review, and editing; JH, Conceptualization, Data curation, Funding acquisition, Writing-review, and editing. All authors critically edited the manuscript, followed by reading and approving the final version. All authors had full access to all the data in the study and had final responsibility for the decision to submit for publication.

## Data availability

The GWAS summary statistic for major depressive disorder from Psychiatric Genomics Consortium (PGC) was available at https://pgc.unc.edu/. GWAS for myocardial infarction was obtained from the GWAS Catalog website (GCST009541). GWAS for all quantitative traits of blood lipids from Global Lipids Genetics Consortium (GLGC) were available at https://csg.sph.umich.edu/willer/public/glgc-lipids2021. The eQTL from GTEx v8 were available at http://gusevlab.org/projects/fusion/. The plasma pQTL from the UK Biobank Pharma Proteomics Project (UKB-PPP) were obtained through the website https://www.synapse.org/, from deCODE genetics via http://www.decode.com, and from ARIC study via http://nilanjanchatterjeelab.org/pwas/. The DLPFC pQTL were obtained at https://www.synapse.org/.

## Code availability

No novel custom computer code or mathematical algorithm was employed in generating the results pivotal to the derived conclusions.

## Acknowledgements

We thank Figdraw (www.figdraw.com) for providing for drawing help. We would like to express our gratitude to all researchers and participants involved in GWAS studies, for their generous sharing their data.

## Reference

1. Murray CJ, Lopez AD. Measuring the global burden of disease. N Engl J Med 2013; 369(5): 448–57.

2. Frasure-Smith N, Lesperance F, Talajic M. Depression following myocardial infarction. Impact on 6-month survival. JAMA 1993; 270(15): 1819–25.

3. Carney RM, Blumenthal JA, Stein PK, et al. Depression, heart rate variability, and acute myocardial infarction. Circulation 2001; 104(17): 2024–8.

4. Kahl KG, Greggersen W, Schweiger U, et al. Prevalence of the metabolic syndrome in unipolar major depression. Eur Arch Psychiatry Clin Neurosci 2012; 262(4): 313–20.

5. Persons JE, Fiedorowicz JG. Depression and serum low-density lipoprotein: A systematic review and meta-analysis. J Affect Disord 2016; 206: 55–67.

6. Oliveri A, Rebernick RJ, Kuppa A, et al. Comprehensive genetic study of the insulin resistance marker TG:HDL-C in the UK Biobank. Nat Genet 2024; 56(2): 212–21.

7. Cross-Disorder Group of the Psychiatric Genomics C. Identification of risk loci with shared effects on five major psychiatric disorders: a genome-wide analysis. Lancet 2013; 381(9875): 1371–9.

8. Turley P, Walters RK, Maghzian O, et al. Multi-trait analysis of genome-wide association summary statistics using MTAG. Nat Genet 2018; 50(2): 229–37.

9. Graham SE, Clarke SL, Wu KH, et al. The power of genetic diversity in genome-wide association studies of lipids. Nature 2021; 600(7890): 675–9.

10. Howard DM, Adams MJ, Clarke TK, et al. Genome-wide meta-analysis of depression identifies 102 independent variants and highlights the importance of the prefrontal brain regions. Nat Neurosci 2019; 22(3): 343–52.

11. Hartiala JA, Han Y, Jia Q, et al. Genome-wide analysis identifies novel susceptibility loci for myocardial infarction. Eur Heart J 2021; 42(9): 919–33.

12. Bulik-Sullivan BK, Loh PR, Finucane HK, et al. LD Score regression distinguishes confounding from polygenicity in genome-wide association studies. Nat Genet 2015; 47(3): 291–5.

13. Ning Z, Pawitan Y, Shen X. High-definition likelihood inference of genetic correlations across human complex traits. Nat Genet 2020; 52(8): 859–64.

14. Werme J, van der Sluis S, Posthuma D, de Leeuw CA. An integrated framework for local genetic correlation analysis. Nat Genet 2022; 54(3): 274–82.

15. Richmond RC, Davey Smith G. Mendelian Randomization: Concepts and Scope. Cold Spring Harb Perspect Med 2022; 12(1).

16. Watanabe K, Taskesen E, van Bochoven A, Posthuma D. Functional mapping and annotation of genetic associations with FUMA. Nat Commun 2017; 8(1): 1826.

17. Foley CN, Staley JR, Breen PG, et al. A fast and efficient colocalization algorithm for identifying shared genetic risk factors across multiple traits. Nat Commun 2021; 12(1): 764.

18. Jiang L, Xue C, Dai S, et al. DESE: estimating driver tissues by selective expression of genes associated with complex diseases or traits. Genome Biol 2019; 20(1): 233.

19. Xue C, Jiang L, Zhou M, et al. PCGA: a comprehensive web server for phenotype-cell-gene association analysis. Nucleic Acids Res 2022; 50(W1): W568–W76.

20. Li M, Jiang L, Mak TSH, et al. A powerful conditional gene-based association approach implicated functionally important genes for schizophrenia. Bioinformatics 2019; 35(4): 628–35.

21. Jiang L, Miao L, Yi G, et al. Powerful and robust inference of complex phenotypes’ causal genes with dependent expression quantitative loci by a median-based Mendelian randomization. Am J Hum Genet 2022; 109(5): 838–56.

22. de Leeuw CA, Mooij JM, Heskes T, Posthuma D. MAGMA: generalized gene-set analysis of GWAS data. PLoS Comput Biol 2015; 11(4): e1004219.

23. Gusev A, Ko A, Shi H, et al. Integrative approaches for large-scale transcriptome-wide association studies. Nat Genet 2016; 48(3): 245–52.

24. Wingo TS, Liu Y, Gerasimov ES, et al. Brain proteome-wide association study implicates novel proteins in depression pathogenesis. Nat Neurosci 2021; 24(6): 810–7.

25. Giambartolomei C, Vukcevic D, Schadt EE, et al. Bayesian test for colocalisation between pairs of genetic association studies using summary statistics. PLoS Genet 2014; 10(5): e1004383.

26. Zhang J, Dutta D, Kottgen A, et al. Plasma proteome analyses in individuals of European and African ancestry identify cis-pQTLs and models for proteome-wide association studies. Nat Genet 2022; 54(5): 593–602.

27. Bennett DA, Buchman AS, Boyle PA, Barnes LL, Wilson RS, Schneider JA. Religious Orders Study and Rush Memory and Aging Project. J Alzheimers Dis 2018; 64(s1): S161–S89.

28. Beach TG, Adler CH, Sue LI, et al. Arizona Study of Aging and Neurodegenerative Disorders and Brain and Body Donation Program. Neuropathology 2015; 35(4): 354–89.

29. Sun BB, Chiou J, Traylor M, et al. Plasma proteomic associations with genetics and health in the UK Biobank. Nature 2023; 622(7982): 329–38.

30. Ferkingstad E, Sulem P, Atlason BA, et al. Large-scale integration of the plasma proteome with genetics and disease. Nat Genet 2021; 53(12): 1712–21.

31. Qi X, Guan F, Wen Y, et al. Integrating genome-wide association study and methylation functional annotation data identified candidate genes and pathways for schizophrenia. Prog Neuropsychopharmacol Biol Psychiatry 2020; 96: 109736.

32. Wu X, Zhang W, Zhao X, et al. Investigating the relationship between depression and breast cancer: observational and genetic analyses. BMC Med 2023; 21(1): 170.

33. Mysling S, Kristensen KK, Larsson M, et al. The angiopoietin-like protein ANGPTL4 catalyzes unfolding of the hydrolase domain in lipoprotein lipase and the endothelial membrane protein GPIHBP1 counteracts this unfolding. Elife 2016; 5.

34. Zhao B, Li T, Fan Z, et al. Heart-brain connections: Phenotypic and genetic insights from magnetic resonance images. Science 2023; 380(6648): abn6598.

35. Morales K, Wittink M, Datto C, et al. Simvastatin causes changes in affective processes in elderly volunteers. J Am Geriatr Soc 2006; 54(1): 70–6.

36. Hyyppa MT, Kronholm E, Virtanen A, Leino A, Jula A. Does simvastatin affect mood and steroid hormone levels in hypercholesterolemic men? A randomized double-blind trial. Psychoneuroendocrinology 2003; 28(2): 181–94.

37. Yang CC, Jick SS, Jick H. Lipid-lowering drugs and the risk of depression and suicidal behavior. Arch Intern Med 2003; 163(16): 1926–32.

38. Otte C, Zhao S, Whooley MA. Statin use and risk of depression in patients with coronary heart disease: longitudinal data from the Heart and Soul Study. J Clin Psychiatry 2012; 73(5): 610–5.

39. Santos M, Mendonca MI, Sa D, et al. ZNF259 rs964184 variant is associated with dyslipidemia and coronary artery disease in the young population. European Heart Journal 2022; 43(Supplement_2).

40. Manthei KA, Yang SM, Baljinnyam B, et al. Molecular basis for activation of lecithin:cholesterol acyltransferase by a compound that increases HDL cholesterol. Elife 2018; 7.

41. Wang Y, Wang S, Zhang L, et al. A simple and precise method to detect sterol esterification activity of lecithin/cholesterol acyltransferase by high-performance liquid chromatography. Anal Bioanal Chem 2018; 410(6): 1785–92.

