## Supplementary Methods for "Role of Blood Lipids in the Shared Genetic Etiology Between Major Depressive Disorder and Myocardial Infarction: A Large-scale Multi-trait Association Analysis"

### 1.Genotypic quality control

Genome-wide association study (GWAS) datasets for major depressive disorder (MDD)[1], myocardial infarction (MI)[2], and 4 quantitative traits of blood lipids (BL) (HDL-C, LDL-C, TC, and TG)[3] used in this study have undergone stringent quality control. These quality control steps were as follows: (i) removing all rare variants with the minor allele frequency (MAF) less than 0.01, (ii) excluding insertions or deletions polymorphisms, (iii) excluding palindromic single nucleotide polymorphisms (SNVs), (iv) removing SNVs within the major histocompatibility complex region (MHC, Chr6: 28,477,797–33,448,354,www.ncbi.nlm.nih.gov/grc/human/regions/MHC?asm=GRCh37) due to its complex linkage disequilibrium (LD) structure, (v) restricting these summary statistics to 1000 Genomes Project Phase 3 European reference SNVs to ensure consistency. In subsequent analyses, corresponding data processing programs were added to adapt for the requirements of different analytical methods.

### 2.Global genetic correlation analysis

We used both linkage disequilibrium (LD) score regression (LDSC)[4] and high-Definition Likelihood (HDL)[5] to evaluate heritability of each trait and genetic correlations between MDD, MI, and the quantitative traits of BL. Following the requirements of the LDSC method, we used pre-computed LD scores from European reference panel in 1000 Genomes Project Phase 3 and retained only about 1.2 million well-imputed HapMap3 SNVs in LDSC analysis (https://github.com/bulik/ldsc)[4]. We did not restrict the intercepts in LDSC analysis, which could account for population stratification or sample overlap. We estimated the heritability of MDD and MI on the liability scale, based on the estimated disease prevalence (6.38%[6] for MDD and 4%[7] for MI). Compared to LDSC only accounting for partial LD information, HDL can fully account for LD across the genome and greatly improve the estimation precision for genetic correlations. We used the 769,306 well-imputed UK Biobank HapMap2 SNVs (https://github.com/zhenin/HDL/wiki/Reference-panels) as reference panel that pre-developed based on HDL approach[5]. For the above statistical analyses, we applied the Benjamini-Hochberg false discovery rate (FDR) approach with a threshold of 0.05 to correct for multiple testing.

**3.Local genetic correlation**

Due to the complex variability of genetic regions, global genetic correlations may potentially underestimate pleiotropic correlations across traits. For example, inconsistencies of the direction on genetic correlations at different regions may have antagonistic effects, which may result in an insignificant overall genetic correlation. Therefore, we additionally used local analysis of [co]variant association (LAVA)[8] to assess local genetic correlation between MDD, MI, and the quantitative traits of BL. We used the genetic covariance intercept which was calculated from the LDSC analysis to correct for potential sample overlap. Taking pre-computed local 2495 genomic locus (https://github.com/josefin-werme/LAVA)[8], we evaluated local genetic correlations across all traits. The results were corrected for multiple testing with FDR method.

**4. Mendelian randomization**

We first performed bidirectional two-sample Mendelian randomization (MR)[9] analyses between MDD and MI. Next, we conducted mediation MR analyses with two-step[10] framework to further explore the role of BL between MDD and MI. For MR analyses, we utilized inverse variance weighted method (IVW)[11,12] as main analysis, and additionally applied several alternative MR methods under different assumptions as sensitivity analyses to further validate the results: (i) MR-Egger[13], which considered the presence of the intercept and can test for pleiotropy; (ii) MR-Radial[14], which can identify single outlying variant that causes large differences between IVW and MR-Egger regression estimates. (iii) MR-

Pleiotropy Residual Sum and Outlier (MRPRESSO)[15], which is able to detect outliers with horizontal pleiotropic effects. We selected the genome-wide significant SNVs with $P < 5 \times 10^{-8}$, and used 1000 Genomes Project phase 3 of European population as LD reference panel to obtain independent instrumental variants with $r^2 < 0.001$ and physical distance > 10,000 kb. However, MR-Radial detected more than 10% outlying variants at the threshold $r^2 < 0.001$, so we set the threshold to a more stringent 0.0001. we used F-statistic to quantify the strength of the instrumental variables[16]. F-statistic larger than 10 indicated the absence of weak bias in the instrumental variable. We utilized steiger test to assess whether the results were affected by potential reverse causality, as well as Cochran's Q test[12] and MR-Egger regression intercept test[13] to respectively assess the presence of heterogeneity and pleiotropy of the results.

In addition, we performed drug target MR[17] analyses to further explored genetically predicted lipid-lowering drug effects on diseases. We included three classes of FDA-approved lipid-lowering drugs: HMGCR inhibitors, PCSK9 inhibitors, and NPC1L1 inhibitors. Of note, according to the parameters as published previously[17], we selected instrumental variants with $r^2 < 0.3$ and physical distance > 100 kb from target gene that was associated with LDL cholesterol level at a genome-wide significance level ($p < 5.0 \times 10^{-8}$) to proxy for lipid-lowering drugs.

**5.Multi-trait association analysis**

Multi-trait analysis of GWAS (MTAG)[18] utilizes a generalized inverse-variance-weighted meta-analysis for multiple correlated traits, which can borrow relevant information from multiple related traits and effectively improve statistical power to identify novel genomic risk loci for each trait. Of note, a key homogeneity assumption of MTAG is that all SNVs share the same variance-covariance matrix. However, the estimator of MTAG is able to be still consistent even if this assumption is violated. The max FDR (the FDR in the worst case) is thus used to evaluate the overall inflation due to violation of the homogeneous assumption. In this study, we performed a MTAG analysis of -$GWAS_{MDD}$, -$GWAS_{MI}$, and -$GWAS_{all\ traits\ of\ BL}$. The genetic correlations between these traits were calculated and further corrected for sample overlap using LDSC method. We calculated the maxFDR to ensure that no overall inflation due to violation of the homogeneous assumption. We set the genome-wide significant threshold at $5\times10^{-8}$.

**6.Functional annotation**

For significant pleiotropic single nucleotide variants (SNVs) from MTAG, we used the Functional Mapping and Annotation of Genetic Associations (FUMA)[19] SNP2GENE function v1.6.1 to characterize significant genomic loci ($P < 5\times10^{-8}$). Using LD information from the 1000 Genome Project phase 3 reference panel of European population, FUMA identified genome-wide significant independent SNVs with $r^2$ less than 0.6 and lead SNVs with $r^2$ less than 0.1 within 1 Mb. Risk loci were defined by combining lead SNVs that physically overlapped or had LD blocks within 250 kb apart. FUMA additionally provided functional annotations such as ANNOVAR (v2017-07-17) analysis, combined annotation dependent depletion (CADD) scores, and RegulomeDB scores. Variant with CADD score exceeding 12.37 were deemed potentially deleterious.

**7.Multi-trait colocalization**

We performed Hypothesis prioritization in multi-trait colocalization (HyPrColoc)[20] analysis to identify pleiotropic genomic risk loci and shared causal variants across the genomic locus of MDD, MI, and the quantitative traits of BL. The HyPrColoc is an extension of the Bayesian colocalization approach, which can effectively colocalize multiple traits simultaneously. We applied R package hyprcoloc (version 1.0) with default settings for analysing, including a prior probability of initial trait association of $1\times10^{-4}$ and a conditional probability of subsequent traits having shared association of 0.02. The HyPrColoc analysis was restricted to the consecutive non-overlapping genetic risk loci for MDD and MI. We declared a colocalized locus when the genomic locus with a posterior probability exceeded 0.75.

**9.Gene-set analysis**

To identify potential biological pathways, we performed MAGMA gene-set[19] enrichment analysis by using the Gene Ontology (GO) and Kyoto Encyclopedia of Genes and Genomes (KEGG) databases. P-values were corrected for multiple testing using FDR correction, and the statistical significance threshold was set at 0.05.

**12.Phenotype-cell-gene association analysis**

Based on the DESE (driver tissue estimation by selective expression)[21] approach, we further explore the associated tissue/cell types and genes for MDD and MI. We implemented in phenotype-cell-gene association analysis (PCGA)[21-24] website to DESE prioritizes disease-driving tissue/cell types by integrating GWAS summary statistics and gene expression profiles, with the underlying hypothesis that disease-associated genes tend to be selectively expressed in disease-driven tissues or cells. Using 54 human tissues (GTEx v8) and 2,214 types of human cells (PanglaoDB, Human Cell Landscape and Allen Brain Atlas), we inspected the enrichment of genes related to MDD and MI.

**8.Gene mapping and gene-based analysis**

Using FUMA v1.5.2 with the default parameters, we performed positional gene-mapping (within a 0kb distance from the locus) analysis to map the genome-wide significant SNVs from loci to specific genes. Next, based on the positional mapping genes, we further performed MAGMA gene-based analysis. The FDR correction was used to control the results for multiple testing.

**10. Transcriptome-wide and colocalization analysis**

We performed transcriptome-wide association studies (TWAS)[25] by using TWAS-Fusion to identify significant gene expression for MDD and MI. We used tissue-specific eQTL data (GTEx v8) from 5 cardiovascular and 13 brain tissues, as well as precomputed functional weights from these eQTL reference panels for analyzing. The results were corrected for multiple testing with FDR method. To confirm that the observed associations did not represent random correlations between gene expression and non-causal variants, we conducted colocalization[26] analysis on the conditionally significant genes ($P < 0.05$) using the default parameters. We declared that if posterior probability for H4 (PP.H4) larger than 0.75, the gene expression and disease association was driven by the shared causal SNV.

**11. Proteome-wide association and colocalization analysis**

Using similar parameters in TWAS-Fusion, we conducted proteome-wide association studies (PWAS)[27] analyses with pQTL from plasma [ARIC study (n = 7,213 European Americans, n = 4,657 proteins)][28] and DLPFC tissues [ROS/MAP study (n = 376 individuals, n = 1,475 proteins)[29] and Banner Sun Health Institute study (n = 152 individuals, n = 1,145 proteins)][30]. We used FDR method to correct the results for multiple testing with FDR method. And we also performed colocalization[26] analyses to estimate the posterior probability of a shared causal variant between the gene expression and diseases association (PP.H4>0.75).

**13. Proteome-wide Mendelian randomization analyses**

We performed proteome-wide MR[11,12] to identify novel potential therapeutic targets for MDD and MI. We selected instrumental variants for each protein in its cis-region (within 1Mb window on either side of gene's transcription site) with $r2 < 0.001$ and physical distance > 10,000 kb. Next, we performed MR analyses using R package Mendelian Randomization (version 0.7.0), taking IVW (fix-IVW for 2-3 instrumental variants, random IVW for over 3 instrumental variants, or Wald ratio (for a single instrumental variant) method as main analysis. The steps of the MR analyses and sensitivity analyses are consistent with the previous description, and the results were corrected for multiple testing with FDR method. This study included three large-scale protein quantitative trait loci (pQTL) summary statistics. Plasma pQTL statistics were obtained from UK Biobank Pharma Proteomics Project (UKB-PPP)[31] and deCODE genetics[32]. The latest published plasma pQTL statistics from the UKB-PPP contains 2,941 plasma proteins in 54,000 UK Biobank participants, of which 34,557 are European ancestry. The pQTL statistics were generated based on Olink Explore 3072. The largest sample size of plasma pQTL statistics based on European ancestry to date was from deCODE (by Ferkingstad, E. et al. in 2021), which included 35,559 Icelandic individuals. A total of 4,907 plasma proteins were measured using the SomaScan multiplex aptamer assay (version 4). Dorsolateral prefrontal cortex (DLPFC) pQTL statistics were obtained from previously described the ROS/MAP study[29].

**14.Bayesian colocalization analysis**

We performed Bayesian colocalization[26] analyses by using the coloc R package (version 5.2.1) to test whether identified associations between proteins and diseases were driven by linkage disequilibrium. based on single causal variant assumption, Bayesian colocalization provided five mutually exclusive hypotheses: H0: there is no genetic association for each trait; H1: there is a genetic association for trait 1 only; H2: there is a genetic association for trait 2 only; H3: there are genetic associations for both traits, but the causal variant for each trait is different; H4: there is a genetic association for both traits, and both traits share the causal variant. Under the default setting, the prior probabilities were set at $1\times10^{-4}$ for P1 when a SNV was associated with trait 1 only, $1\times10^{-4}$ for P2 when a SNV was associated with trait 2 only, and $1\times10^{-5}$ for P12 when a SNV as associated with both traits. We declared that when the posterior probability for H4 (PP.H4) exceeded 0.75, the genetic association were driven by the shared causal SNV.

**Reference**

1. Howard DM, Adams MJ, Clarke TK, et al. Genome-wide meta-analysis of depression identifies 102 independent variants and highlights the importance of the prefrontal brain regions. *Nat Neurosci* 2019; **22**(3): 343-52.

2. Hartiala JA, Han Y, Jia Q, et al. Genome-wide analysis identifies novel susceptibility loci for myocardial infarction. *Eur Heart J* 2021; **42**(9): 919-33.

3. Graham SE, Clarke SL, Wu KH, et al. The power of genetic diversity in genome-wide association studies of lipids. *Nature* 2021; **600**(7890): 675-9.

4. Bulik-Sullivan BK, Loh PR, Finucane HK, et al. LD Score regression distinguishes confounding from polygenicity in genome-wide association studies. *Nat Genet* 2015; **47**(3): 291-5.

5. Ning Z, Pawitan Y, Shen X. High-definition likelihood inference of genetic correlations across human complex traits. *Nat Genet* 2020; **52**(8): 859-64.

6. Arias-de la Torre J, Vilagut G, Ronaldson A, et al. Prevalence and variability of current depressive disorder in 27 European countries: a population-based study. *Lancet Public Health* 2021; **6**(10): e729-e38.

7. Benjamin EJ, Virani SS, Callaway CW, et al. Heart Disease and Stroke Statistics-2018 Update: A Report From the American Heart Association. *Circulation* 2018; **137**(12): e67-e492.

8. Werme J, van der Sluis S, Posthuma D, de Leeuw CA. An integrated framework for local genetic correlation analysis. *Nat Genet* 2022; **54**(3): 274-82.

9. Richmond RC, Davey Smith G. Mendelian Randomization: Concepts and Scope. *Cold Spring Harb Perspect Med* 2022; **12**(1).

10. Carter AR, Sanderson E, Hammerton G, et al. Mendelian randomisation for mediation analysis:

current methods and challenges for implementation. *Eur J Epidemiol* 2021; **36**(5): 465-78.

11. Burgess S, Scott RA, Timpson NJ, Davey Smith G, Thompson SG, Consortium E-I. Using published data in Mendelian randomization: a blueprint for efficient identification of causal risk factors. *Eur J Epidemiol* 2015; **30**(7): 543-52.

12. Burgess S, Butterworth A, Thompson SG. Mendelian randomization analysis with multiple genetic variants using summarized data. *Genet Epidemiol* 2013; **37**(7): 658-65.

13. Bowden J, Davey Smith G, Burgess S. Mendelian randomization with invalid instruments: effect estimation and bias detection through Egger regression. *Int J Epidemiol* 2015; **44**(2): 512-25.

14. Bowden J, Spiller W, Del Greco MF, et al. Improving the visualization, interpretation and analysis of two-sample summary data Mendelian randomization via the Radial plot and Radial regression. *Int J Epidemiol* 2018; **47**(6): 2100.

15. Verbanck M, Chen CY, Neale B, Do R. Detection of widespread horizontal pleiotropy in causal relationships inferred from Mendelian randomization between complex traits and diseases. *Nat Genet* 2018; **50**(5): 693-8.

16. Palmer TM, Lawlor DA, Harbord RM, et al. Using multiple genetic variants as instrumental variables for modifiable risk factors. *Stat Methods Med Res* 2012; **21**(3): 223-42.

17. Huang W, Xiao J, Ji J, Chen L. Association of lipid-lowering drugs with COVID-19 outcomes from a Mendelian randomization study. *Elife* 2021; **10**.

18. Turley P, Walters RK, Maghzian O, et al. Multi-trait analysis of genome-wide association summary statistics using MTAG. *Nat Genet* 2018; **50**(2): 229-37.

19. Watanabe K, Taskesen E, van Bochoven A, Posthuma D. Functional mapping and annotation of genetic associations with FUMA. *Nat Commun* 2017; **8**(1): 1826.

20. Foley CN, Staley JR, Breen PG, et al. A fast and efficient colocalization algorithm for identifying shared genetic risk factors across multiple traits. *Nat Commun* 2021; **12**(1): 764.

21. Jiang L, Xue C, Dai S, et al. DESE: estimating driver tissues by selective expression of genes associated with complex diseases or traits. *Genome Biol* 2019; **20**(1): 233.

22. Xue C, Jiang L, Zhou M, et al. PCGA: a comprehensive web server for phenotype-cell-gene association analysis. *Nucleic Acids Res* 2022; **50**(W1): W568-W76.

23. Li M, Jiang L, Mak TSH, et al. A powerful conditional gene-based association approach implicated functionally important genes for schizophrenia. *Bioinformatics* 2019; **35**(4): 628-35.

24. Jiang L, Miao L, Yi G, et al. Powerful and robust inference of complex phenotypes' causal genes with dependent expression quantitative loci by a median-based Mendelian randomization. *Am J Hum Genet* 2022; **109**(5): 838-56.

25. Gusev A, Ko A, Shi H, et al. Integrative approaches for large-scale transcriptome-wide association studies. *Nat Genet* 2016; **48**(3): 245-52.

26. Giambartolomei C, Vukcevic D, Schadt EE, et al. Bayesian test for colocalisation between pairs of genetic association studies using summary statistics. *PLoS Genet* 2014; **10**(5): e1004383.

27. Wingo TS, Liu Y, Gerasimov ES, et al. Brain proteome-wide association study implicates novel proteins in depression pathogenesis. *Nat Neurosci* 2021; **24**(6): 810-7.

28. Zhang J, Dutta D, Kottgen A, et al. Plasma proteome analyses in individuals of European and African ancestry identify cis-pQTLs and models for proteome-wide association studies. *Nat Genet* 2022; **54**(5): 593-602.

29. Bennett DA, Buchman AS, Boyle PA, Barnes LL, Wilson RS, Schneider JA. Religious Orders Study and Rush Memory and Aging Project. *J Alzheimers Dis* 2018; **64**(s1): S161-S89.

30. Beach TG, Adler CH, Sue LI, et al. Arizona Study of Aging and Neurodegenerative Disorders and Brain and Body Donation Program. *Neuropathology* 2015; **35**(4): 354-89.

31. Sun BB, Chiou J, Traylor M, et al. Plasma proteomic associations with genetics and health in the UK Biobank. *Nature* 2023; **622**(7982): 329-38.

32. Ferkingstad E, Sulem P, Atlason BA, et al. Large-scale integration of the plasma proteome with genetics and disease. *Nat Genet* 2021; **53**(12): 1712-21.
