## Supplementary Figure 1-3 for "Role of Blood Lipids in the Shared Genetic Etiology Between Major Depressive Disorder and Myocardial Infarction: A Large-scale Multi-trait Association Analysis"

**Supplementary Fig. 1 Significantly enriched pathways for major depressive disorder and myocardial infarction in GO/KEGG enrichment analysis.** a, enriched pathways for major depressive disorder. b, enriched pathways for myocardial infarction. *GO: Gene Ontology, MF: molecular function, CC: cellular component, BP: biological process, KEGG: Kyoto Encyclopedia of Genes and Genomes.*

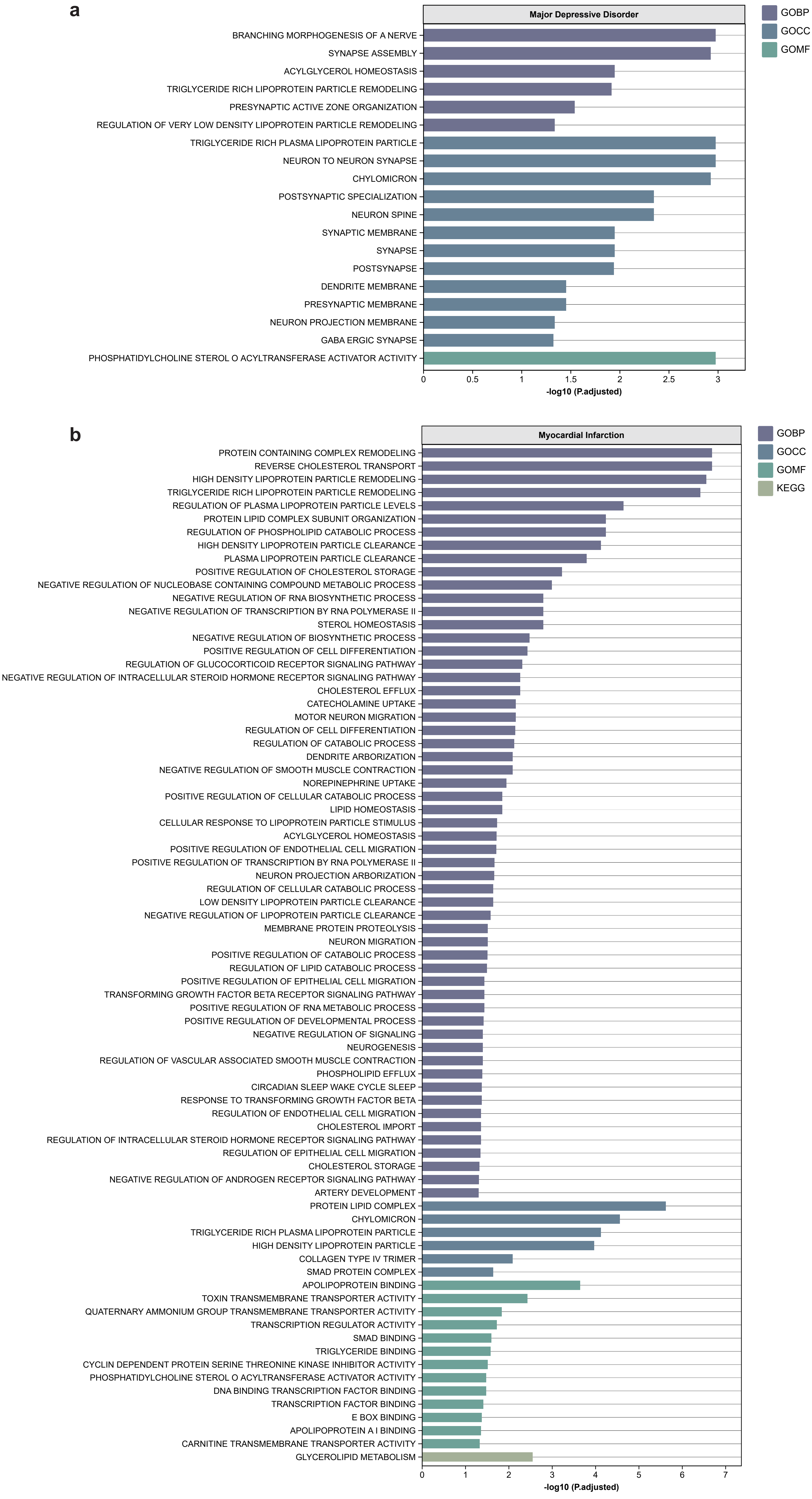

**Supplementary Fig. 2 Tissue specificity association for major depressive disorder and myocardial infarction.** a, associated tissues for major depressive disorder. b, associated tissues for myocardial infarction.

a

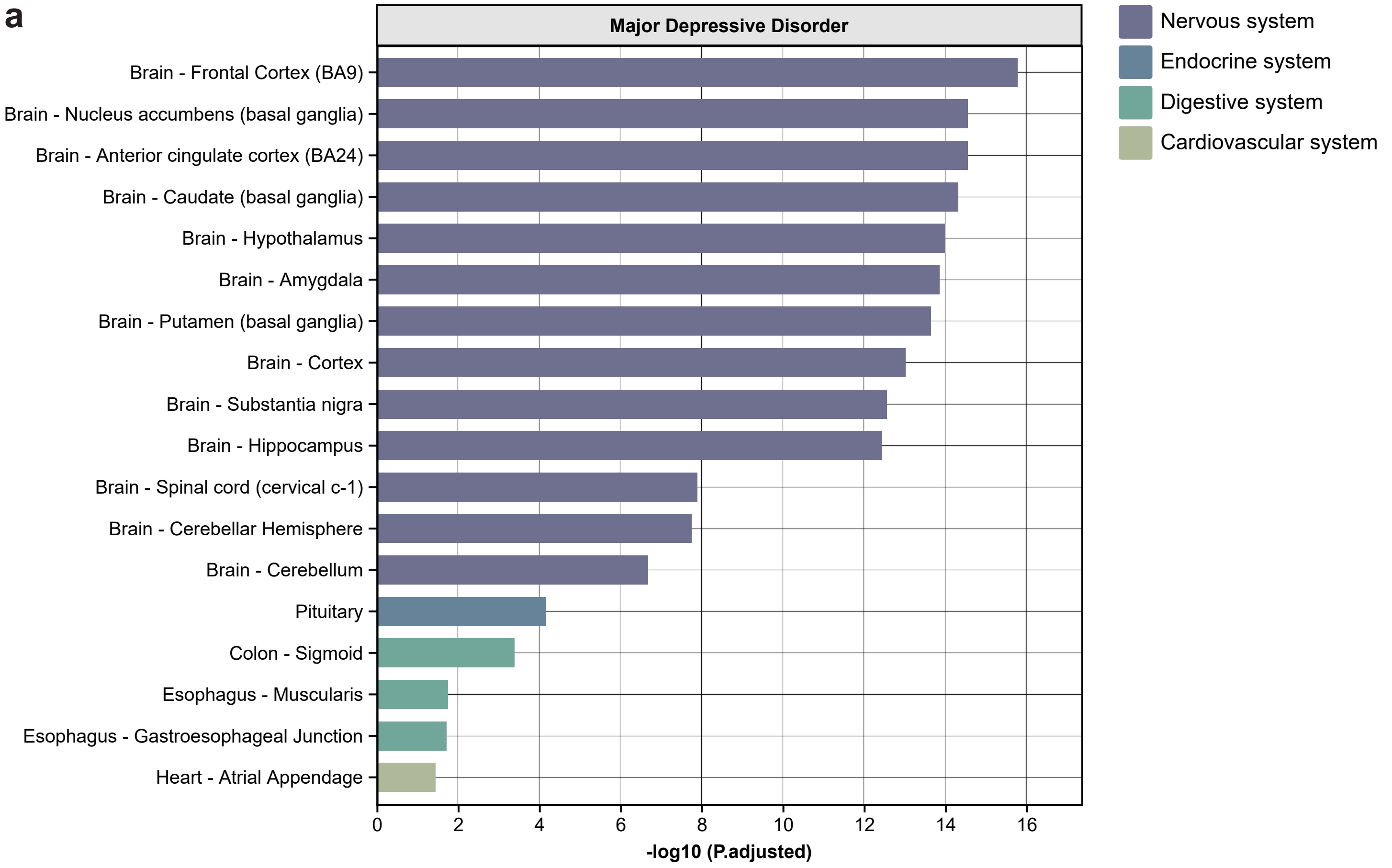


b

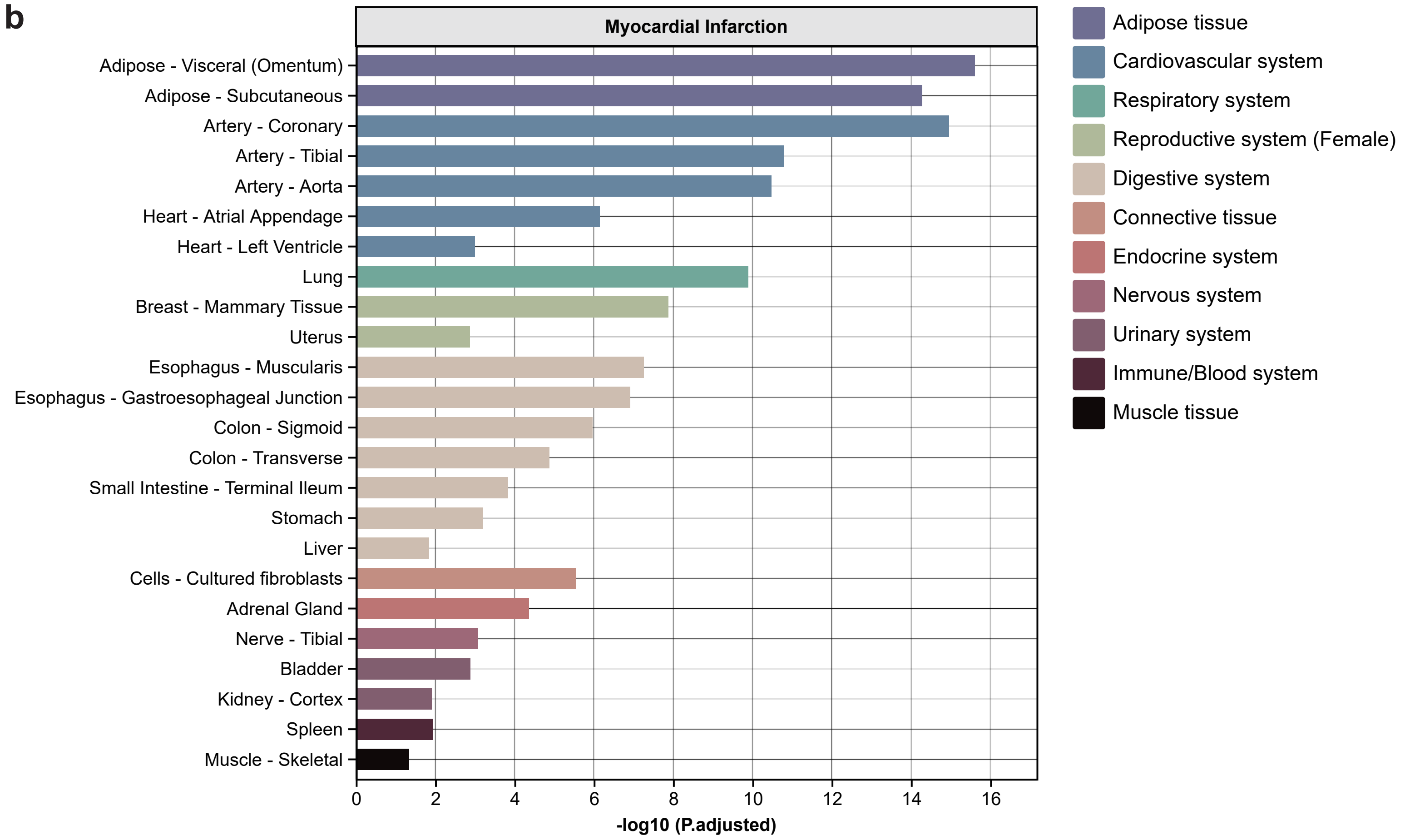

**Supplementary Fig. 3 Cell specificity association for major depressive disorder and myocardial infarction.** a, associated cell types for major depressive disorder. b, associated cell types for myocardial infarction.

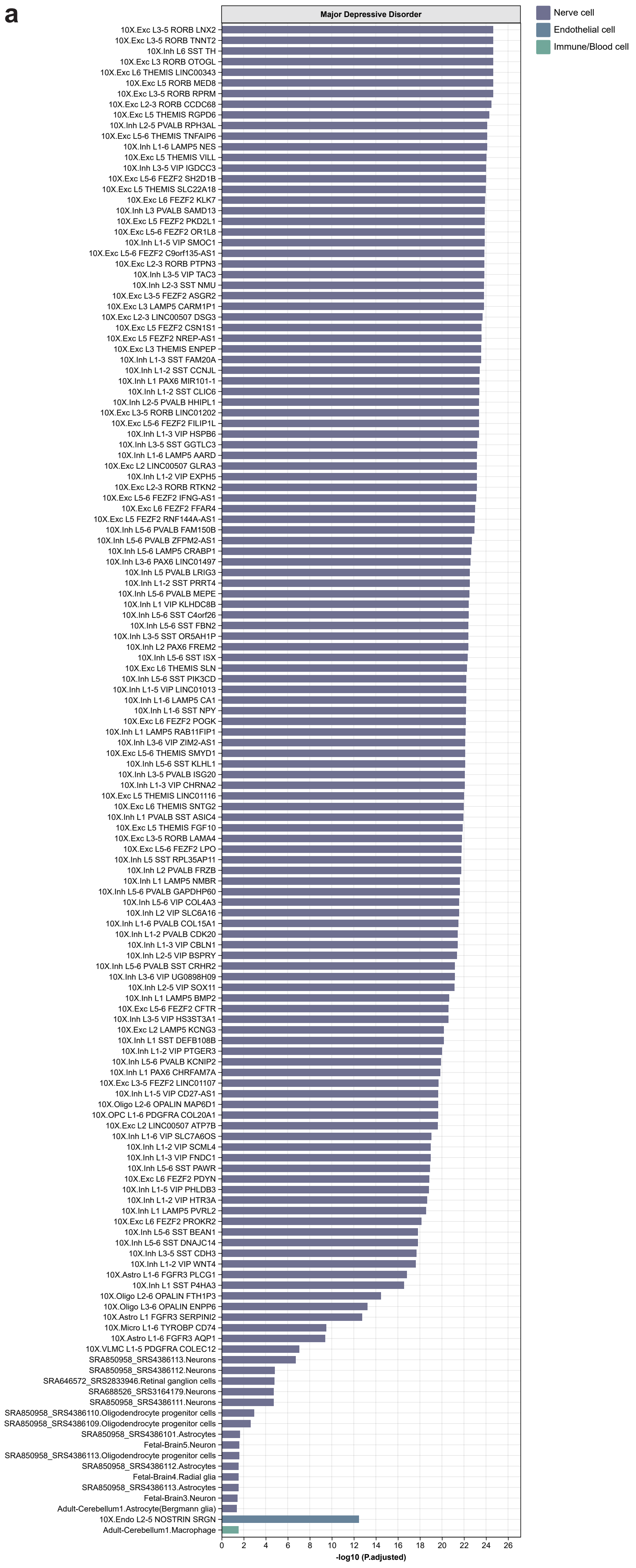

b

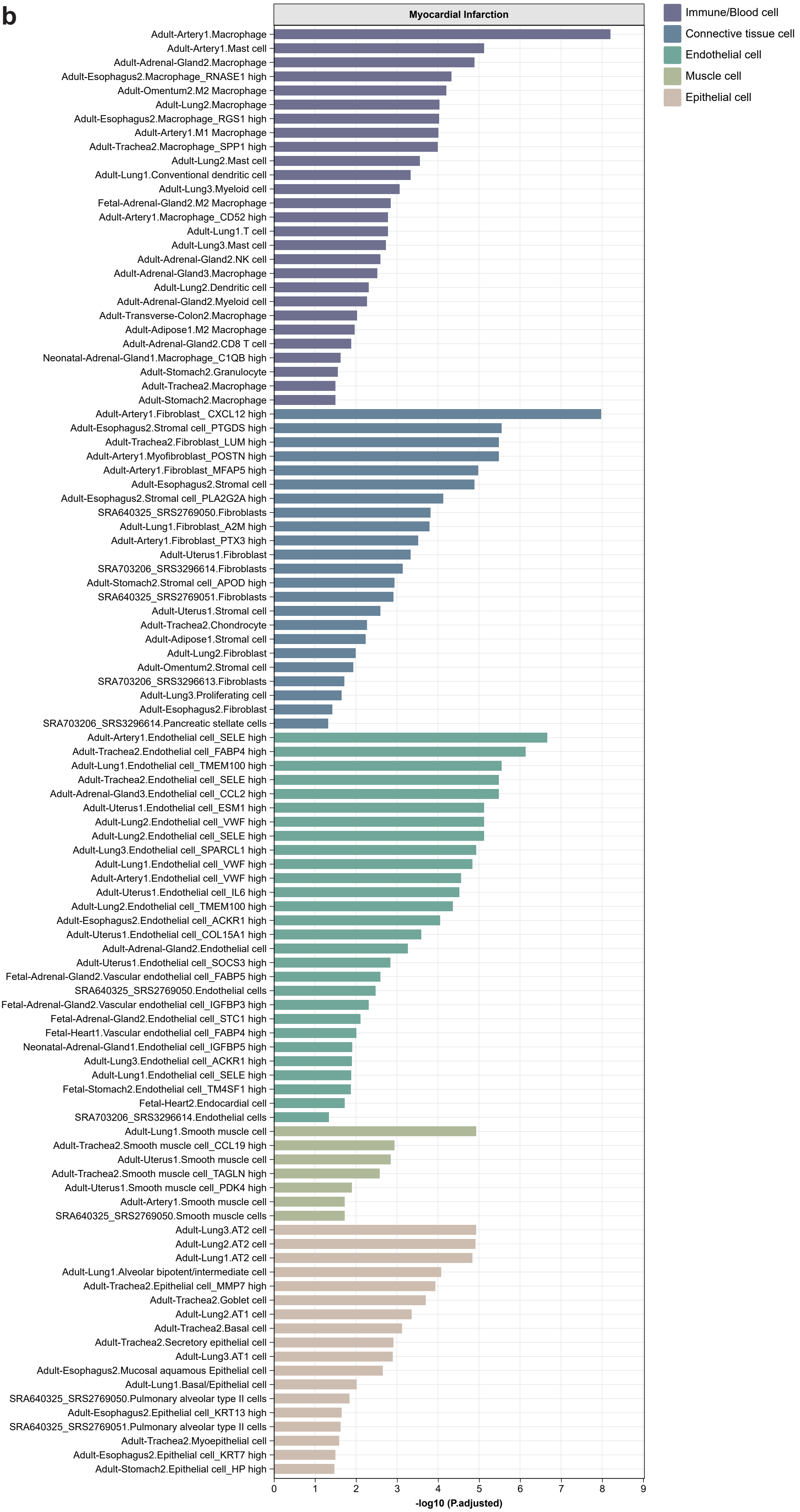
Myocardial Infarction
Immune/Blood cell
Connective tissue cell
Endothelial cell
Muscle cell
Epithelial cell
Adult-Artery1.Macrophage
Adult-Artery1.Mast cell
Adult-Adrenal-Gland2.Macrophage
Adult-Esophagus2.Macrophage_RNASE1 high
Adult-Omentum2.M2 Macrophage
Adult-Lung2.Macrophage
Adult-Esophagus2.Macrophage_RGS1 high
Adult-Artery1.M1 Macrophage
Adult-Trachea2.Macrophage_SPP1 high
Adult-Lung2.Mast cell
Adult-Lung1.Conventional dendritic cell
Adult-Lung3.Myeloid cell
Fetal-Adrenal-Gland2.M2 Macrophage
Adult-Artery1.Macrophage_CD52 high
Adult-Lung1.T cell
Adult-Lung3.Mast cell
Adult-Adrenal-Gland2.NK cell
Adult-Adrenal-Gland3.Macrophage
Adult-Lung2.Dendritic cell
Adult-Adrenal-Gland2.Myeloid cell
Adult-Transverse-Colon2.Macrophage
Adult-Adipose1.M2 Macrophage
Adult-Adrenal-Gland2.CD8 T cell
Neonatal-Adrenal-Gland1.Macrophage_C1QB high
Adult-Stomach2.Granulocyte
Adult-Trachea2.Macrophage
Adult-Stomach2.Macrophage
Adult-Artery1.Fibroblast_ CXCL12 high
Adult-Esophagus2.Stromal cell_PTGDS high
Adult-Trachea2.Fibroblast_LUM high
Adult-Artery1.Myofibroblast_POSTN high
Adult-Artery1.Fibroblast_MFAP5 high
Adult-Esophagus2.Stromal cell
Adult-Esophagus2.Stromal cell_PLA2G2A high
SRA640325_SRS2769050.Fibroblasts
Adult-Lung1.Fibroblast_A2M high
Adult-Artery1.Fibroblast_PTX3 high
Adult-Uterus1.Fibroblast
SRA703206_SRS3296614.Fibroblasts
Adult-Stomach2.Stromal cell_APOD high
SRA640325_SRS2769051.Fibroblasts
Adult-Uterus1.Stromal cell
Adult-Trachea2.Chondrocyte
Adult-Adipose1.Stromal cell
Adult-Lung2.Fibroblast
Adult-Omentum2.Stromal cell
SRA703206_SRS3296613.Fibroblasts
Adult-Lung3.Proliferating cell
Adult-Esophagus2.Fibroblast
SRA703206_SRS3296614.Pancreatic stellate cells
Adult-Artery1.Endothelial cell_SELE high
Adult-Trachea2.Endothelial cell_FABP4 high
Adult-Lung1.Endothelial cell_TMEM100 high
Adult-Trachea2.Endothelial cell_SELE high
Adult-Adrenal-Gland3.Endothelial cell_CCL2 high
Adult-Uterus1.Endothelial cell_ESM1 high
Adult-Lung2.Endothelial cell_VWF high
Adult-Lung2.Endothelial cell_SELE high
Adult-Lung3.Endothelial cell_SPARCL1 high
Adult-Lung1.Endothelial cell_VWF high
Adult-Artery1.Endothelial cell_VWF high
Adult-Uterus1.Endothelial cell_IL6 high
Adult-Lung2.Endothelial cell_TMEM100 high
Adult-Esophagus2.Endothelial cell_ACKR1 high
Adult-Uterus1.Endothelial cell_COL15A1 high
Adult-Adrenal-Gland2.Endothelial cell
Adult-Uterus1.Endothelial cell_SOCS3 high
Fetal-Adrenal-Gland2.Vascular endothelial cell_FABP5 high
SRA640325_SRS2769050.Endothelial cells
Fetal-Adrenal-Gland2.Vascular endothelial cell_IGFBP3 high
Fetal-Adrenal-Gland2.Endothelial cell_STC1 high
Fetal-Heart1.Vascular endothelial cell_FABP4 high
Neonatal-Adrenal-Gland1.Endothelial cell_IGFBP5 high
Adult-Lung3.Endothelial cell_ACKR1 high
Adult-Lung1.Endothelial cell_SELE high
Fetal-Stomach2.Endothelial cell_TM4SF1 high
Fetal-Heart2.Endocardial cell
SRA703206_SRS3296614.Endothelial cells
Adult-Lung1.Smooth muscle cell
Adult-Trachea2.Smooth muscle cell_CCL19 high
Adult-Uterus1.Smooth muscle cell
Adult-Trachea2.Smooth muscle cell_TAGLN high
Adult-Uterus1.Smooth muscle cell_PDK4 high
Adult-Artery1.Smooth muscle cell
SRA640325_SRS2769050.Smooth muscle cells
Adult-Lung3.AT2 cell
Adult-Lung2.AT2 cell
Adult-Lung1.AT2 cell
Adult-Lung1.Alveolar bipotent/intermediate cell
Adult-Trachea2.Epithelial cell_MMP7 high
Adult-Trachea2.Goblet cell
Adult-Lung2.AT1 cell
Adult-Trachea2.Basal cell
Adult-Trachea2.Secretory epithelial cell
Adult-Lung3.AT1 cell
Adult-Esophagus2.Mucosal aquamous Epithelial cell
Adult-Lung1.Basal/Epithelial cell
SRA640325_SRS2769050.Pulmonary alveolar type II cells
Adult-Esophagus2.Epithelial cell_KRT13 high
SRA640325_SRS2769051.Pulmonary alveolar type II cells
Adult-Trachea2.Myoepithelial cell
Adult-Esophagus2.Epithelial cell_KRT7 high
Adult-Stomach2.Epithelial cell_HP high
0
1
2
3
4
5
6
7
8
9
-log10 (P.adjusted)
